# Molecular subtypes of respiratory Adenovirus infection outbreak in children in Northern Vietnam and risk factors of more severe cases

**DOI:** 10.1101/2023.04.18.23288722

**Authors:** Dinh-Dung Nguyen, Lan Tuyet Phung, Huyen Thi Thanh Tran, Ha Thi Thanh Ly, Anh Hang Mai Vo, Nhung Phuong Dinh, Phuong Mai Doan, Anh Thi Nguyen, Luc Danh Dang, Thia Thi Doan, Khuong Thi Pham, Huong Lan Pham, Dai Hoang Xuan, Thao Phuong Nguyen, Bao Thai Tran, Trang Thi Thuc Tran, Huong Thi Minh Le, An Nhat Pham, Antony Antoniou, Nhan Thi Ho

## Abstract

**Background:** Under the pressure of the outbreak of respiratory Human Adenovirus (HAdV) infections in children in Northern Vietnam in the end of 2022, this study was initiated to identify the HAdV subtype(s) responsible for the outbreak in relation to the clinical features of the patients and examine the risk factors of more severe cases.

**Methods:** The study was conducted on pediatric patients tested positive with HAdV using multiplex real-time PCR between October and November 2022. Nasal swab samples were used for sequencing to identify HAdV subtypes and clinical data were collected retrospectively.

**Results:** Among 97 successfully sequenced samples, the predominant subtypes were HAdV-B3 (83%), HAdV-B7 (16%) and HAdV-C2 (1%). Lower respiratory manifestations were found in 25% of patients (5% with severe pneumonia). There was no significant association between HAdV type and clinical features except that those infected with HAdV type 3 exhibited higher WBC and neutrophil % (p<0.001). Co-infection of HAdV with ≥1 other respiratory viruses or bacteria was found in 70.8% of those with lower respiratory illnesses (OR (95%CI); p-value vs. those without =5.21 (1.60, 19.36); 0.0084 after adjusting for age at hospital visit, sex, birth delivery method, day of disease at hospital visit), and in 100% of those with severe pneumonia vs. 33% of those without (p=0.005).

**Conclusion:** HAdV-B3 and HAdV-B7 were predominant in the outbreak. Co-infection of HAdV together with other respiratory viruses or bacteria was a strong risk factor for lower respiratory tract illnesses and severe pneumonia. The findings advocate the advantages of multi-factor microbial panels for the diagnosis and prognosis of respiratory infections in children.

## Introduction

Human Adenoviruses (HAdV) can cause many common diseases such as upper respiratory infection, conjunctivitis, gastrointestinal disorders, etc. HAdV consists of many serotypes, which are responsible for different disease contexts (1).

There had been some sporadic HAdV outbreaks with respiratory illnesses, conjunctivitis, and gastrointestinal disorders worldwide before the COVID-19 era. Respiratory illnesses were reported in most of these studies, and HAdV infections with severe respiratory illnesses in children had been reported in some studies (3–26). The most prevalent respiratory HAdV reported in children in Taiwan were HAdV- B genotype 3 and HAdV-C (HAdV-2), and 7 (15,24,26), in China were HAdV-B3 and HAdV-C2, and 7 (9,10,12,17,27), in Japan were HAdV type 3, 2, and 1 (21), in Argentina were HAdV-B7 and HAdV-B3 (16,20), in Spain were HAdV-3, HAdV-6, and HAdV-5 (25), in Egypt were type 3 and 7 (28), in Palestine were HAdV-C1, HAdV-C2, HAdV-B3 and HAdV-C5 (6). HAdV type 7, 3, and 2 were the most common types which were associated with severe respiratory illnesses in children, especially in those with underlying diseases or immunocompromised conditions (10–12,16–19). During the early stages of the COVID-19 pandemic with social distancing until 2021, respiratory tract infections in children including HAdV infections exhibited a remarkable decrease in occurrence (14,29–31).

In Vietnam, there have been some reports of adenoviral conjunctivitis (32,33), and gastroenteritis (13,34,35). HAdV is among the less common viral causes of seasonal respiratory infections as compared to many other more common viruses such as influenza, parainfluenza, respiratory syncytial virus (RSVs), enteroviruses, rhinovirus, etc (36–43). HAdV is among the co-infection factors in patients with severe pneumonia (39). HAdV pneumonia especially type 7 has been reported to be among the causes of death in patients following measles infection (44,45). There has not been any information regarding respiratory HAdV outbreaks in children in Vietnam until the first half of 2022.

Since the second half of 2022, HAdV had emerged as the cause of the outbreak of respiratory infections in children in Northern Vietnam. Whilst most of the cases were mild, there had been a considerable number of cases with severe complications such as pneumonia, acute respiratory distress. which required intensive treatment and prolonged hospitalization. In addition, quite a few deaths attributed to HAdV infection had been reported. While the COVID-19 pandemic had been somewhat under control, the outbreak of respiratory HAdV infection in children in Northern Vietnam with a large number of cases and an alarming number of severe cases and deaths was unprecedented and thus especially worrisome and required immediate actions.

Up till now, in Northern Vietnam, the diagnosis of HAdV infection was confirmed by positive PCR of nasal swabs performed by a few centers. However, molecular typing of HAdV had not been performed to identify the HAdV subtype(s) responsible for the outbreak, especially in severe or fatal cases. As such, this study aimed to perform molecular typing of HAdV and examine the relationship between HAdV types and clinical contexts of children with respiratory HAdV infections in Northern Vietnam to understand the characteristics of the pathogen in relation to host characteristics. The study also examined potential risk factors for the severity of the diseases to help orientate the treatment and prognosis of the patients.

## Materials & Methods

### Sample Collection

This study was done in Vinmec Times City International Hospital, a private general hospital in Hanoi Vietnam which receives patients mostly from Hanoi and surrounding provinces. The study was approved by the hospital’s ethical committee before being carried out (Reference number: 152/2022/CN-HDDD VMEC).

Nasopharyngeal wash or swab samples of pediatric inpatients and outpatients with respiratory symptoms suspected of Adenoviral infection were collected in a 15.0 ml centrifuge tube and transported to the Department of Microbiology. Specimens were stored at 4°C and partly used to extract RNA by QIAamp Viral RNA Kits (QIAGEN, German) within 24 hours after collection. Extracted RNA samples were tested for HadV and 6 other respiratory pathogens (human enterovirus (HEV), human metapneumovirus (hMPV) and parainfluenza virus 1-4 (PIV1-4)) using multiplex real-time PCR Allplex™ Respiratory Panel 2 (Avicalab Diagnostics, Kenya) according to the manufacturer’s instruction (46,47). Samples with quantification cycle (Ct) value < 38 were considered HAdV-positive.

The leftover nasopharyngeal samples (NS) of those with HAdV-positive PCR were stored and transported to Vinmec Medical Genetic Department for further processing to perform HAdV molecular typing. To ensure a sufficient DNA concentration for sequencing and to minimize possible artifacts due to contamination, only samples with Ct for HAdV <30 were selected for molecular typing.

For pediatric patients with HAdV-positive PCR, clinical data regarding medical history especially prior to COVID infection, clinical manifestations, lab tests (hematology, biochemistry and also nasal-pharyngeal swab culture, PCR panel for 7 respiratory bacteria, blood *Mycoplasma pneumoniae* Antibody test, influenza antigen rapid test if available), treatments, outcomes, etc were retrospectively collected by Vinmec Pediatric Center. These data were entered and managed in RedCap (https://www.project-redcap.org/).

All patients included in this study had respiratory symptoms such as cough, runny nose, etc. Patients were classified as having lower respiratory illnesses if having lower respiratory symptoms such as rales, rhonchi, wheezing, etc or having lesions in chest x-ray. Patients were classified as having severe pneumonia if having signs of pneumonia clinically and/or in chest x-ray and respiratory distress.

Viral co-infection was defined if a patient with HAdV-positive PCR was also positive with at least one virus in the PCR panel or in the influenza antigen rapid test.

Bacterial co-infection was defined if a patient with HAdV-positive PCR was also positive with at least one bacteria in nasopharyngeal fluid culture or in a PCR panel of 7 respiratory bacteria (*H. influenzae, S. pneumonia, S. aereus, M. catarrhalis, M. pneumoniae, C. pneumoniae, L. pneumoniae*) or in blood *Mycoplasma pneumoniae* antibody test.

Any co-infection was defined if a patient had either a viral or bacterial co-infection as described above.

### HAdV molecular typing

The hyper-variable regions 1-6 (HRV_1-6_) of HadV exon are known to contain type-specific epitopes, therefore, these sequences are commonly used to identify HAdV subtypes responsible for the diseases. HAdV molecular typing was performed on 97 HAdV-positive samples. Viral nucleic acids were extracted from NS using QIAamp DNA Blood Mini Kit (QIAGEN, Germany). Following the protocol recommended by CDC, the *hexon* of HadV was amplified using nested PCR targeting the HVR_1-6_ and amplicons were expected to have the size between 764-896 base pairs (bp) (48). The outer primer pairs used for 1st round of nested-PCR were AdhexF1 (5′- TICTTTGACATICGIGGIGTICTIGA-3′) and AdhexR1 (5′- CTGTCIACIGCCTGRTTCCACA-3′). The inner primer pairs used for 2nd round of nested-PCR were AdhexF2 (5′- GGYCCYAGYTTYAARCCCTAYTC-3′) and AdhexR2 (5′- GGTTCTGTC ICCCAGAGARTCIAGCA-3′). Nested-PCR aws performed using a final concentration of 1x GoTaq Green Master Mix (Promega), 0.4 μM of each primer, 2.0 μl of extracted viral nucleic acid or 0.2 μl of 1st nested-PCR product, and made up to a final volume of 25 μl with double distilled water. The PCR thermal program for both rounds of nested-PCR reactions were comprised an initial denaturation step at 94°C for 2 min, followed by 11 touched-down cycles of 95°C for 30 sec, 55°C (decrease 1°C after each cycle) for 45 sec, 72°C for 1 min, followed by 20 thermal cycles of 95°C for 30 sec, 45°C for 45 sec, 72°C for 1 min, and a 5 min final extension at 72°C. The size of PCR products were checked by DNA electrophoresis analysis on 1% agarose gels stained with RedSafe DNA Stain (20,000 X) (Chembio, USA). Only samples with a detectable appropriate PCR product from nested-PCR were used for Sanger Sequencing of the HVR_1-6_ of hexon gene.

Each nested-PCR product was sequenced using both forward AdhexF2 and reverse AdhexR2 primers. Sequencing PCR reactions were performed in a final volume of 10 μl comprising 0.5 μl of BigDye Terminator v3.1 Cycle Sequencing (ABI), 2.0 μl of Sequencing Buffer 5X, 0.85 μl (10 μM) of primer, 0.2μl 2nd nested-PCR product, and distilled water (up-to-10 μl). The sequencing thermal cycle for sequencing reactions were comprised of an initial denaturation step at 96°C for 2 min, followed by 25 thermal cycle of 95°C for 10 sec, 50°C for 10 sec, and 60°C for 2 min. Cycle sequencing products were purified by Bigdye X Terminator purification kit before analysis on th Applied Biosystems 3500 Dx Genetic Analyzer.

### Data analysis

#### Sequence data and phylogenetic analysis

The sequence data were used to identify the subtype of HAdV by Basic Local Alignment Search Tool (BLAST) search (NCBI). The sequences of HVR_1-6_ of *hexon* gene from all samples were aligned with related reference strains obtained from Genbank (reference strains are listed in Supplementary Table 1) using ClustalW. The robustness of the phylogenetic tree was constructed by the neighbor-joining method with bootstrap analysis (n = 1000) by iTOL v6 and Adobe Illustrator software. The cut-off value is <80%.

#### Analysis of HAdV types and clinical contexts

HAdV types and other clinical features were compared between disease contexts such as lower respiratory illnesses vs. no lower respiratory illnesses or severe pneumonia vs no severe pneumonia. The purpose was to examine if HAdV type or any other clinical features were associated with the severity of HAdV infection. Clinical features were also compared between HAdV types to examine if any clinical characteristic was associated with HAdV types.

For comparison between groups for initial association exploration, Kruskall-Wallis was used for continuous variables and Fisher’s exact test was used for categorical variables.

Potential risk factors for lower respiratory illnesses or severe pneumonia which were clinically relevant or statistically significant from the above exploration were further examined using univariate and multivariate logistic regression models. Unadjusted and adjusted odds ratio (OR) and 95% confidence interval (95%CI) by profile likelihood and p-values from Wald test were reported.

Data analysis was done using R statistical software version 4.1 (https://www.r-project.org). All statistical tests were 2-sided with a significant level alpha of 0.05.

## Results

### Patient characteristics and clinical features

From October 13th to November 9th, 2022, 138 patients tested positive for HAdV using a Q-PCR assay from nasal swabs. Among these, 97 patients with Ct for HAdV <30 were selected for molecular typing. Of these 97 samples, the median (IQR) Ct of HAdV was 23.6 (21.9, 25.6).

Patient characteristics overall and comparison between those with and without lower respiratory illnesses and those with and without severe pneumonia are summarized in Table 1. About 68% of patients resided in Hanoi and 32% resided in surrounding provinces. About 29% reported history of prior COVID-19 infection. About 4% reported history of allergy and about 7% reported history of prior severe respiratory diseases. There were 73 (75%) patients with only upper respiratory symptoms whilst there were 24 (25%) with lower respiratory manifestations of which 5 (5.1%) had severe pneumonia. There were no differences in demographic and previous history of respiratory disease including COVID19 infection between those with and without lower respiratory illnesses in these characteristics (Table 1, Figure 1, Figure 2).

**Figure 1.**
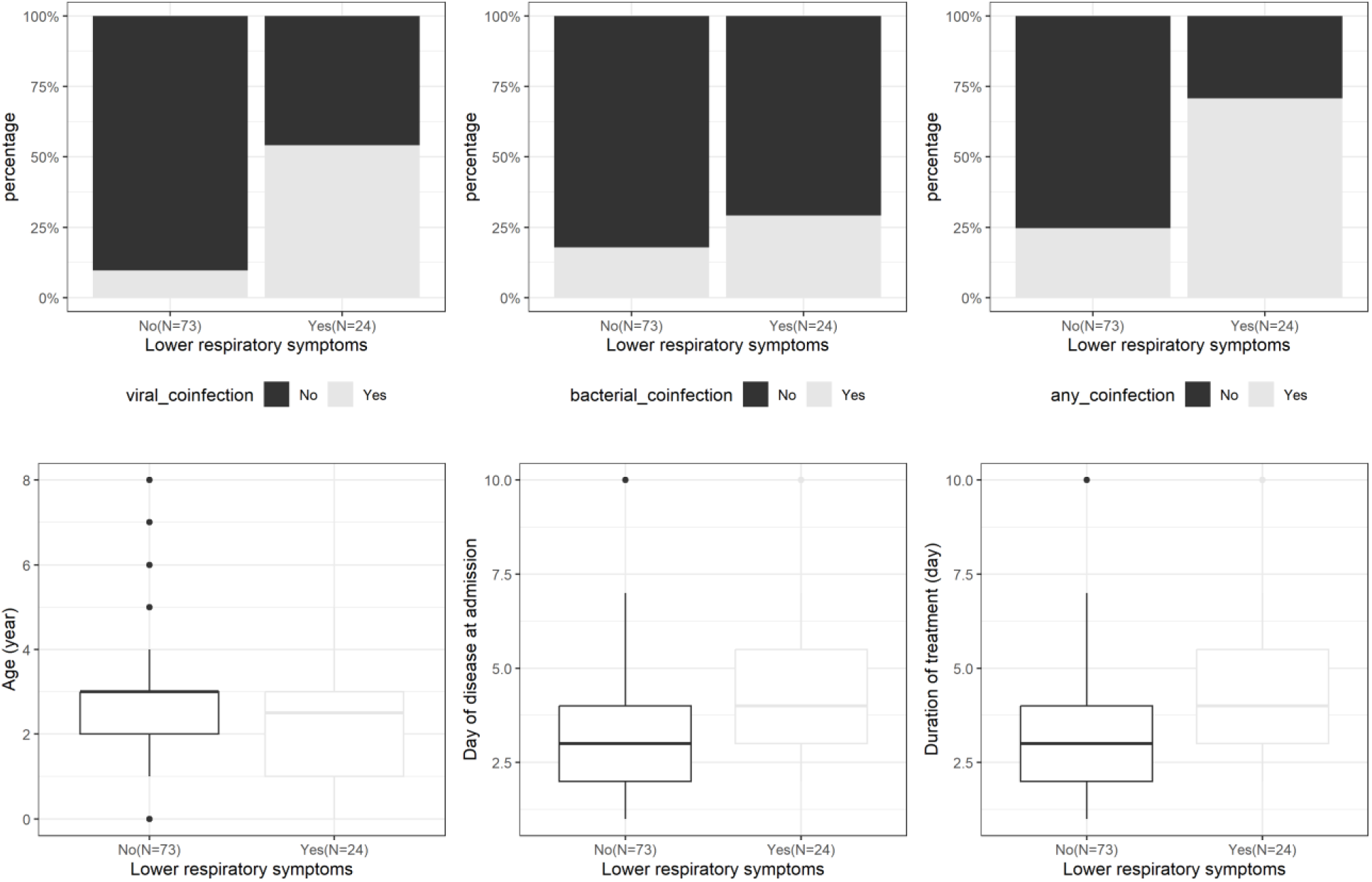
Patient characteristics by lower respiratory illnesses. Upper panel: co-infection of human Adenovirus (HAdV) with other viruses or bacteria in patients with lower respiratory illnesses vs. those without. Lower panel: some clinical features of patients with lower respiratory illnesses vs. those without. Viral_coinfection: co-infection of HAdV with ≥1 other viruses in PCR panel or influenza virus type A or type B using rapid antigen test Vacterial_coinfection: co-infection of HAdV with ≥1 bacteria via nasopharyngeal fluid culture or PCR panel of 7 respiratory bacteria (H. influenzae, S. pneumonia, S. aereus, M. catarrhalis, M. pneumoniae, C. pneumoniae, L. pneumoniae) or Mycoplasma antibody test. Any_coinfection: co-infection of HAdV with ≥1 bacteria or viruses mentioned above.

**Figure 2.**
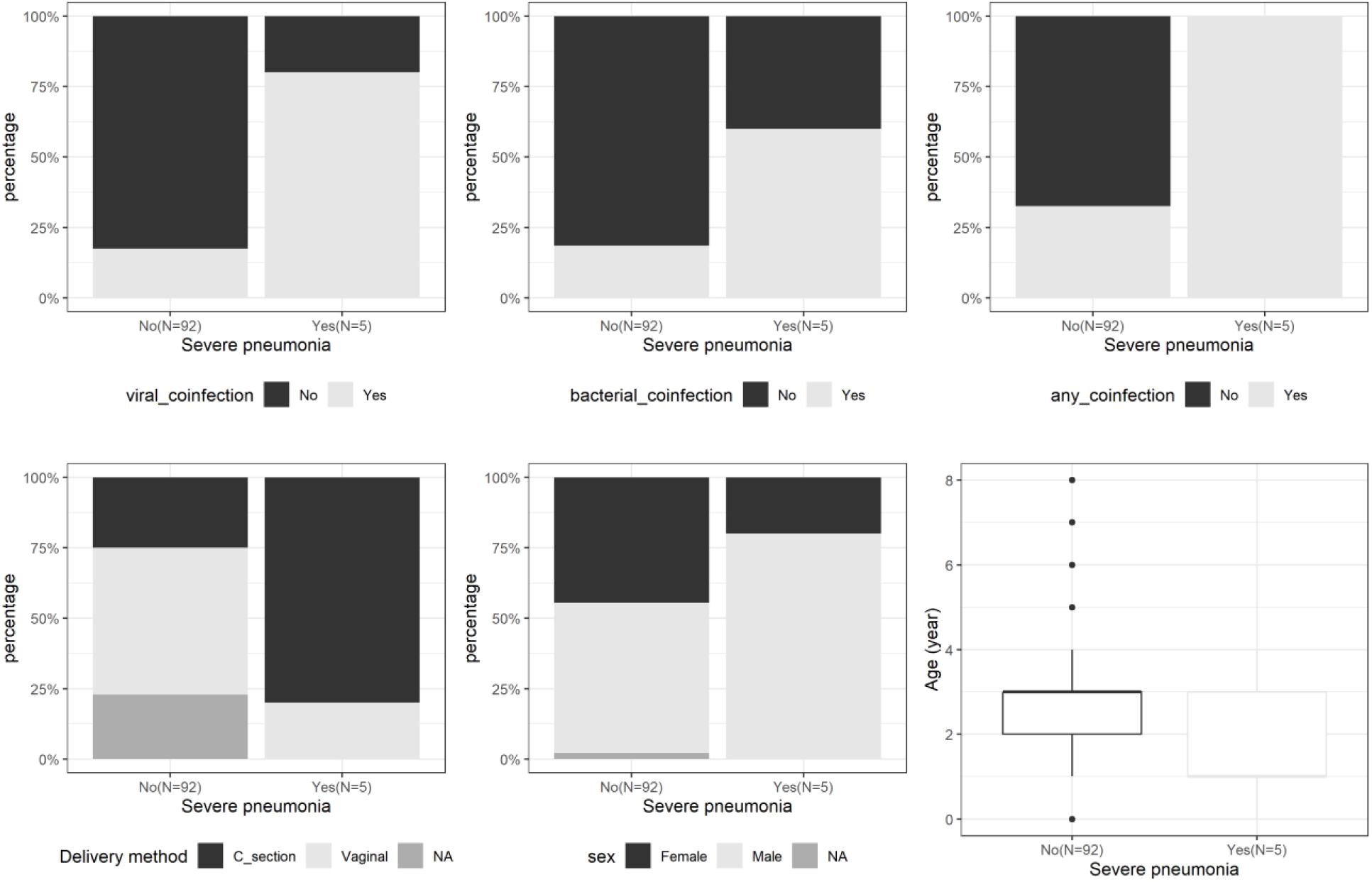
Patient characteristics by severe pneumonia. Upper panel: co-infection of human Adenovirus (HAdV) with other viruses or bacteria in patients with severe pneumonia vs. those without. Lower panel: some clinical features of patients with severe pneumonia vs. those without. viral_coinfection: co-infection of HAdV with ≥1 other viruses in PCR panel or influenza virus type A or type B using rapid antigen test bacterial_coinfection: co-infection of HAdV with ≥1 bacteria via nasopharyngeal fluid culture or PCR panel of 7 respiratory bacteria (H. influenzae, S. pneumonia, S. aereus, M. catarrhalis, M. pneumoniae, C. pneumoniae, L. pneumoniae) or Mycoplasma antibody test. any_coinfection: co-infection of HAdV with ≥1 bacteria or viruses mentioned above.

**Table 1.** Patient characteristics overall and in groups with lower respiratory illnesses and severe pneumonia.

| Characteristics | All patients (N=97) | Group with lower respiratory illnesses (N=24) | P-value comparison* | Group with severe pneumonia (N=5) | P-value comparison # |
| --- | --- | --- | --- | --- | --- |
| HAdV type | Type 3= 81(84%);<br>Type 7= 15 (15%);<br>Type 2= 1 (1%) | Type 3= 19 (79.2%);<br>Type 7= 5 (20.8%);<br>Type 2=0 | 0.636 | Type 3= 4 (80.0%);<br>Type 7= 1 (20.0%);<br>Type 2=0 | 0.602 |
| Viral co-infection\$ | 20 (20.6%) | 12 (50.0%) | < 0.001 | 4 (80.0%) | 0.006 |
| Bacterial co-infection @ | 20 (20.6%) | 7 (29.2%) | 0.253 | 3 (60.0%) | 0.058 |
| Any co-infection \$@ | 35 (36.1%) | 17 (70.8%) | < 0.001 | 5 (100.0%) | 0.005 |
| Sex (males) | 53 (55.8%) | 13 (54.2%) | 1 | 4 (80.0%) | 0.379 |
| Age at hospital visit (mean (95%CI)) | 2.7 (2.4, 3.1) | 2.3 (1.6, 2.9) | 0.141 | 1.8 (0, 3.8) | 0.190 |
| BMI (kg/m2) (mean (95%CI)) | 15.3 (14.9, 15.6) | 15 (14.2, 15.9) | 0.497 | 14.7 (13.7, 15.7) | 0.487 |
| Gestational age (week) (mean (95%CI)) | 37.6 (36.8, 38.5) | 37.8 (36.9, 38.7) | 0.517 | 36 (36, 36) | 0.115 |
| Delivery method | C-section= 27 (35.5%);<br>Vaginal= 49 (64.5%) | C-section= 11 (50.0%);<br>Vaginal= 11 (50.0%) | 0.116 | C-section= 4 (80.0%);<br>Vaginal= 1 (20.0%) | 0.051 |
| Breast feeding duration (month) (mean (95%CI)) | 12.1 (10.9, 13.3) | 13.2 (10.8, 15.6) | 0.410 | 12.3 (2.7, 21.8) | 0.575 |
| History of COVID-19 infection | 18 (29.0%) | 4 (26.7%) | 1 | 1 (50.0%) | 0.500 |
| History of allergy | 4 (4.3%) | 1 (4.2%) | 1 | 0 (0.0%) | 1 |
| History of immuno-compromise diseases | 0 | 0 | NA | 0 | NA |
| History of chronic disease | 0 | 0 | NA | 0 | NA |
| History of severe respiratory disease | 7 (7.2%) | 1 (4.2%) | 0.677 | 1 (20.0%) | 0.318 |
| Day of disease at hospital visit (mean (95%CI)) | 3.6 (3.2, 4) | 4.7 (3.7, 5.6) | 0.002 | 4 (2, 6) | 0.422 |
| Cough | 88 (95.7%) | 23 (95.8%) | 1 | 4 (80.0%) | 0.203 |
| Fever | 88 (90.7%) | 21 (87.5%) | 0.685 | 5 (100.0%) | 1.000 |
| Duration of fever (day) (mean (95%CI)) | 6.2 (5.7, 6.8) | 7.1 (5.7, 8.6) | 0.125 | 6.2 (1.8, 10.6) | 0.839 |
| Red eye | 30 (33.3%) | 5 (21.7%) | 0.207 | 0 (0.0%) | 0.165 |
| Loose stool | 15 (16.7%) | 6 (25.0%) | 0.352 | 1 (20.0%) | 1 |
| Lymph node enlargement | 6 (6.7%) | 1 (4.2%) | 1 | 0 (0.0%) | 1 |
| Skin rash | 1 (1.1%) | 0 (0.0%) | 1 | 0 (0.0%) | 1 |
| Coughing up sputum | 86 (93.5%) | 22 (91.7%) | 1 | 5 (100.0%) | 1 |
| Ear pain | 5 (5.9%) | 1 (4.3%) | 0.653 | 0 (0.0%) | 1 |
| Abdominal pain | 7 (7.9%) | 2 (8.7%) | 1 | 0 (0.0%) | 1 |
| Poor eating | 38 (42.2%) | 16 (66.7%) | 0.007 | 4 (80.0%) | 0.158 |
| WBC (mean (95%CI)) | 15 (13.5, 16.5) | 13.8 (10.9, 16.7) | 0.295 | 13.5 (3.6, 23.5) | 0.661 |
| Neutrophil % (mean (95%CI)) | 61.2 (58, 64.4) | 59.5 (52.4, 66.6) | 0.659 | 64.5 (41.9, 87) | 0.496 |
| Lymphocyte % (mean (95%CI)) | 26.8 (23.8, 30) | 29.9 (22.9, 36.9) | 0.405 | 26.8 (4.8, 48.9) | 0.766 |
| Hemoglobin (g/L) (mean (95%CI)) | 85.5 (74.7, 96.3) | 84.6 (64.2, 105) | 0.364 | 89.1 (34.5, 143.7) | 0.538 |
| Platelet (mean (95%CI)) | 295 (275, 315) | 304 (250, 357) | 0.811 | 412 (183, 641) | 0.104 |
| CRP (mean (95%CI)) | 40.4 (32.8, 48.1) | 41.3 (22.3, 60.3) | 0.773 | 37.2 (0.5, 73.9) | 0.949 |
| Chest X-ray abnormality | 22 (51.2%) | 19 (95.0%) | < 0.001 | 5 (100.0%) | 0.048 |
| Duration of treatment (day) (mean (95%CI)) | 5 (4.5, 5.6) | 6.1 (4.7, 7.4) | 0.020 | 9.5 (4.9, 14.1) | 0.003 |
| Oxy supplement | 6 (6.2%) | 6 (25.0%) | <0.001 | 4 (80.0%) | < 0.001 |
| Intravenous Immunoglobulin (IVIG) infusion for Adenovirus treatment | 2 (2.9%) | 2 (9.5%) | 0.098 | 2 (40.0%) | 0.004 |
| Nebulizer | 10 (14.3%) | 10 (43.5%) | < 0.001 | 4 (80.0%) | 0.001 |
| Corticoid use | 5 (7.4%) | 5 (23.8%) | 0.002 | 4 (80.0%) | < 0.001 |
| Antibiotic use | 78 (86.7%) | 23 (95.8%) | 0.170 | 5 (100.0%) | 1.000 |
| Clinical outcome | Dead=0 (0.0%);<br>Discharged with complication =1 (1.2%);<br>Discharged without | Dead=0 (0.0%);<br>Discharged with complication = 1 (4.8%);<br>Discharged without | 0.6941 | Dead=0 (0.0%);<br>Discharged with complication = 1 (20.0%);<br>Discharged without | 0.0630 |
|  | complication<br>= 82 (98.8%) | complication<br>= 20 (95.2%) |  | complication<br>= 4 (80.0%) |  |

\*Comparison between those with and without lower respiratory illnesses.
#Comparison between those with and without severe pneumonia.
P-values from Kruskal-Wallis test for continuous variables or Fisher's exact test for categorical variables.

The mean (95%CI) age of all patients was 2.7 (2.4, 3) years and those with lower respiratory illnesses were a bit younger than those without (mean (95%CI) = 2.3 (1.6, 2.9) vs. 3.1 (2.6, 3.6) respectively) and those with severe pneumonia were the youngest (mean (95%CI) = 1.8 (0, 3.8)). Males accounted for 55% of all patients and there were no gender bias in those with and without lower respiratory illnesses whereas 4/5 (80%) of those with severe pneumonia were males. About 43% of patients were born by Caesaren (C)- section and the percentage was similar between those with and without lower respiratory illnesses while 4/5 (80%) of those with severe pneumonia were born by C_section (p=0.051). There was no difference in BMI, gestational age, birth weight and duration of breastfeeding between those with and without lower respiratory illnesses or between those with and without severe pneumonia (Table 1).

Day of illness at hospital visit of those with lower respiratory illnesses was later than those without (mean (95%CI) = 4.7 (3.7, 5.6) days vs. 3.3 (2.8, 3.8)) days (p=0.002)). Most of the patients had fever (94%) and cough (84%) at disease onset. Most of the patients (79%) had high fever (>39 C) and long fever duration (mean (95%CI) = 6.4 (5.7, 7.1) days). Poor eating (as compared to usual) as reported by caregivers was found more in patients with lower respiratory illnesses than those without. There was no significant difference in symptoms at disease onset such as cough, red eyes, diarrhea, fever maximal temperature, fever duration, ear pain, etc as well as in hematology or biochemistry indexes between those with and without lower respiratory illnesses or between those with and without severe pneumonia. Almost all patients with lower respiratory illnesses (95%) had detectable abnormalities in chest X-rays as well as 16% of those without lower respiratory symptoms (Table 1, Figure 1 and Figure 2).

About 25% of the patients with lower respiratory illnesses received oxygen supplements, about 10% received IVIG, 44% received nebulizers, 24% received general steroids while none of the patients without lower respiratory illnesses received these interventions. Duration of hospitalization was longer for those with lower respiratory illnesses than those without (mean (95%CI) = 6.1 (4.7, 7.4) vs. 4.5 (3.8, 5.1) days) and longest in those with severe pneumonia (mean (95%CI) = 9.5 (4.9, 14.1) days). All patients except one were discharged without complication (Table 1, Figure 1 and 2).

The five patients with severe pneumonia were of a younger age and were predominantly male (80%), of which 80% were born by C-section and were all co-infected with at least one other respiratory viruses or bacteria (p<0.05 as compared to other patients). There was no other remarkable difference in other demographic, medical history or host characteristics as compared to other patients.

### Co-infection of HAdV and other respiratory viruses or bacteria

Regarding viral co-infection, there were 19 (approx. 20%) samples tested positive for at least one other virus from our respiratory virus PCR panel of which 5(5.1%) were co-infected with at least 2 viruses. The number of samples co-infected with HEV, MPV, PIV1, PIV2, PIV3, PIV4 in the PCR panel were 7, 9, 4, 2, 0, 2 respectively. There were two samples co-infected with Influenza virus detected by rapid antigen test. In overall, there were 20 samples co-infected with at least 1 virus detected by either PCR or rapid antigen test. Regarding bacterial co-infection, there were 3, 11, 5 samples with positive culture for Streptococcus pneumoniae, Haemophilus influenzae, Moraxella catarrhalis respectively. There were 2 patients with blood antibody test positive for Mycoplasma pneumoniae. In overall, there were 20 patients co-infected with at least one bacterium detected by either nasopharyngeal fluid culture or blood antibody test. There were 5 patients co-infected with both at least one virus and at least one bacterium. In total, there were 35 patients co-infected with either virus or bacteria (Table 2).

**Table 2.** Viral and bacterial co-infection in addition to human Adenovirus (HAdV).

| **Viral co-infection** | **Any virus** | **HEV\*** | **MPV\*** | **PIV1\*** | **PIV2\*** | **PIV3\*** | **PIV\*4** | **Influenza#** |
| --- | --- | --- | --- | --- | --- | --- | --- | --- |
| Number of patients | 20 | 7 | 9 | 4 | 2 | 0 | 2 | 1 |

| **Bacterial co-infection** | **Any bacteria** | **Streptococcus pneumoniae\$** | **Haemophilus influenzae** | **Moraxella catarrhalis** | **Mycoplasma pneumoniae** | **Others** | | |
| --- | --- | --- | --- | --- | --- | --- | --- | --- |
| Number of patients | 20 | 3 | 11 | 5 | 2 | 0 |  |  |
| **Viral and bacterial co-infection** | **Any virus and bacteria** |  |  |  |  |  |  |  |
| Number of patients | 5 |  |  |  |  |  |  |  |
| **Either viral or bacterial co-infection** | **Any virus or bacteria** |  |  |  |  |  |  |  |
| Number of patients | 35 |  |  |  |  |  |  |  |

\*Viruses in the same PCR panel with human Adenovirus for nasal swab: HEV: human enterovirus, hMPV: human metapneumovirus, PIV1-4: parainfluenza virus 1-4.
#By rapid antigen test for nasal swab

The percentage of samples co-infected with at least one other respiratory virus was significantly higher in patients with lower respiratory illnesses vs. those without (12 (50%) vs. 4 (9%), p< 0.001) and especially higher in those with severe pneumonia (4/5 (80%), p=0.006). Co-infection with at least one respiratory bacterium was detected in 20 (20.6%) patients, with 7 of these patients (29.2%) experiencing lower respiratory illnesses (p=0.253) and in 3 (60.0%) with severe pneumonia (p=0.058). Any co-infection with respiratory viruses or bacteria was found in 35 (36.1%) of all patients, significantly higher in those with lower respiratory illnesses (17 (70.8%), p< 0.001) and especially higher in those with severe pneumonia (5 (100.0%), p=0.005) (Table 1, Table 2).

### Risk factors for lower respiratory illnesses and severe pneumonia

Potential risk factors for lower respiratory illnesses or severe pneumonia which were clinically relevant and statistically significant from the above exploration were viral co-infection of HAdV with ≥1 respiratory viruses or bacterial co-infection of HAdV with ≥1 respiratory bacteria or any co-infection of HAdV with ≥1 respiratory viruses or bacteria, age at hospital visit, sex, birth delivery method, day of disease at hospital visit. The results from univariate logistic regression and multivariate logistic regression adjusting for these factors in the models for the comparison between those with lower respiratory illnesses or severe pneumonia vs. those without are summarized in Table 3. Co-infection of HAdV with ≥1 other respiratory viruses was remarkably significantly associated with either lower respiratory illnesses or severe pneumonia (unadjusted OR (95%CI); (p-value) for those with lower respiratory illnesses or severe pneumonia vs. those without were 11.14 (3.77, 36.05); <0.0001 and 19.00 (2.60, 385.24); 0.0105 respectively). After adjusting for age at hospital visit, sex, birth delivery method, day of disease at hospital visit, the association between co-infection and lower respiratory illnesses or severe pneumonia remained statistically significant (adjusted OR (95%CI); (p-value) for those with lower respiratory illnesses or severe pneumonia vs. those without were 7.4 (1.4, 52.1); 0.03 and 16.9 (1.8, 421.9); 0.03 respectively). Co-infection of HAdV with any respiratory viruses or bacteria was also remarkably significantly associated with lower respiratory illnesses (unadjusted OR (95%CI); (p-value) = 7.42 (2.75, 21.98); 0.0001 and adjusted OR (95%CI); (p-value) = 5.21 (1.60, 19.36); 0.0084).

**Table 3.** Univariate and multivariate logistic regression of risk factors of lower respiratory illnesses or severe pneumonia.

|  | Lower respiratory illnesses |  | Severe pneumonia |  |
| --- | --- | --- | --- | --- |
| Features | Univariate OR (95%CI); p-value* | Multivariate OR (95%CI); p-value** | Univariate OR (95%CI); p-value# | Multivariate OR (95%CI); p-value## |
| Viral co-infection\$<br>(yes vs. no) | 11.14 (3.77,<br>36.05); <0.0001 | 10.74 (2.83,<br>48.17); 0.0009 | 19.00 (2.60,<br>385.24); 0.0105 | 19.44 (2.12,<br>492.73); 0.0207 |
| Bacterial co-infection (yes vs.<br>no)@ | 1.90 (0.63,<br>5.44); 0.2373 | 1.04 (0.26,<br>3.83); 0.9506 | 6.62 (1.02,<br>53.28); 0.0470 | 19.25 (1.74,<br>528.71); 0.0297 |
| Any co-infection<br>(yes vs. no)\$@ | 7.42 (2.75,<br>21.98); 0.0001 | 5.21 (1.60,<br>19.36); 0.0084 | Inf (NA,NA); NA<br>### | Inf (NA,NA); NA<br>### |
| Age (year) | 0.71 (0.48,<br>1.00); 0.0678 | 0.72 (0.41,<br>1.18); 0.2175 | 0.56 (0.24,<br>1.13); 0.1462 | 0.86 (0.32,<br>2.21); 0.7466 |
| Sex (male vs.<br>female) | 0.92 (0.36,<br>2.35); 0.8531 | 0.52 (0.13,<br>1.86); 0.3226 | 3.35 (0.47,<br>66.86); 0.2884 | 1.46 (0.11,<br>37.77); 0.7823 |
| Delivery method<br>(vaginal vs. C-section) | 0.42 (0.15,<br>1.17); 0.0964 | 0.45 (0.12,<br>1.63); 0.2235 | 0.12 (0.01,<br>0.87); 0.0642 | 0.11 (0.00,<br>1.14); 0.1002 |
| Day of disease at<br>hospital visit | 1.46 (1.13,<br>1.95); 0.0060 | 1.48 (1.09,<br>2.13); 0.0204 | 1.10 (0.66,<br>1.63); 0.6483 | 1.21 (0.65,<br>2.10); 0.4884 |

\*Comparison between those with and without lower respiratory illnesses. 95%CI from profile likelihood and p-values from Wald test of univariate logistic regression model against each variable.
\*\*Comparison between those with and without lower respiratory illnesses. 95%CI from profile likelihood and p-values from Wald test for each variable in the multivariate logistic regression model. For viral co-infection or bacterial co-infection or any co-infection, estimates are from the multivariate logistic regression model containing either viral co-infection or bacterial co-infection or any co-infection and 4 other variables. For 4 other variables (age, sex, delivery method, day of disease at hospital visit), estimates are from the multivariate logistic regression model containing viral co-infection and these 4 variables..

### HAdV types and the relationship between HAdV types and clinical features

Ninety-seven (98.98%) HAdV samples were genotyped successfully. BLAST results of 97 HAdV sequences indicated that 82 (84.5%) samples belonged to subtype B3, 14 (14.4%) samples belonged to subtype C7 and 1 (1.0%) samples belonged to sybtype C2. The phylogenetic analysis were applied to compare 97 hexon gene sequences to each other and 24 reference sequences downloaded from GenBank (Supplement Table 1). The phylogenetic tree of hexon gene confirmed the hexon sequences of HAdV samples belonged to three distinguishable clusters of HAdV-B3, -B7 and C2 (Figure 3).

**Figure 3.**
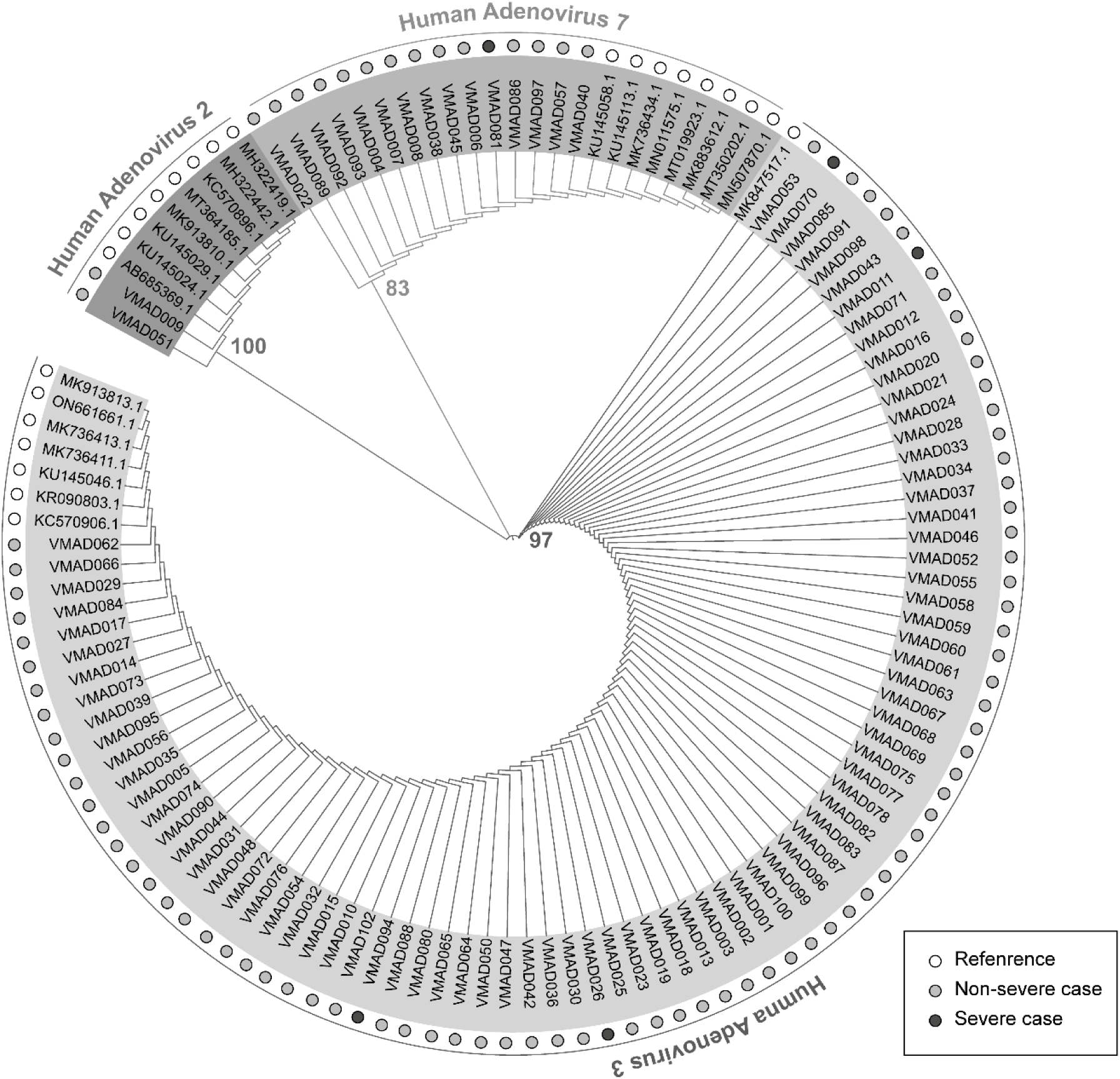
Adenovirus types and phylogenetic tree. Neighbour-joining phylogenetic of the *hexon* gene of HAdV strains identified in patients hospitalized with respiratory tract infections in Vinmec Times City Hospital between October and November 2022. The branch annotations represent the bootstrap value calculated on 1000 replicates. White, green and red circles are corresponding to sequences of HAdV references, HAdV specimens from non-severe patients and HAdV specimens from patients with severe pneumonia. Those infected with HAdV type 3 had higher WBC, higher neutrophil percentage and lower lymphocyte percentage (mean (95%CI) of WBC, neutrophil %, lymphocyte % of those infected with HAdV type 3 vs. type 7 were 16.2 (14.6, 17.8) vs. 9.4 (7.1, 11.7), 64.8 (61.6, 68) vs. 45.6 (39.2, 51.9), 23.3 (20.5, 26.2) vs. 41.9 (34.8, 49) respectively) (p<0.001). In general, there was no remarkable difference in demographic, medical history, disease manifestation or outcome features between patients infected with HAdV type 3, 7, 2 in this study except a significant difference in hematology test mentioned above (Figure 4).

**Figure 4.**
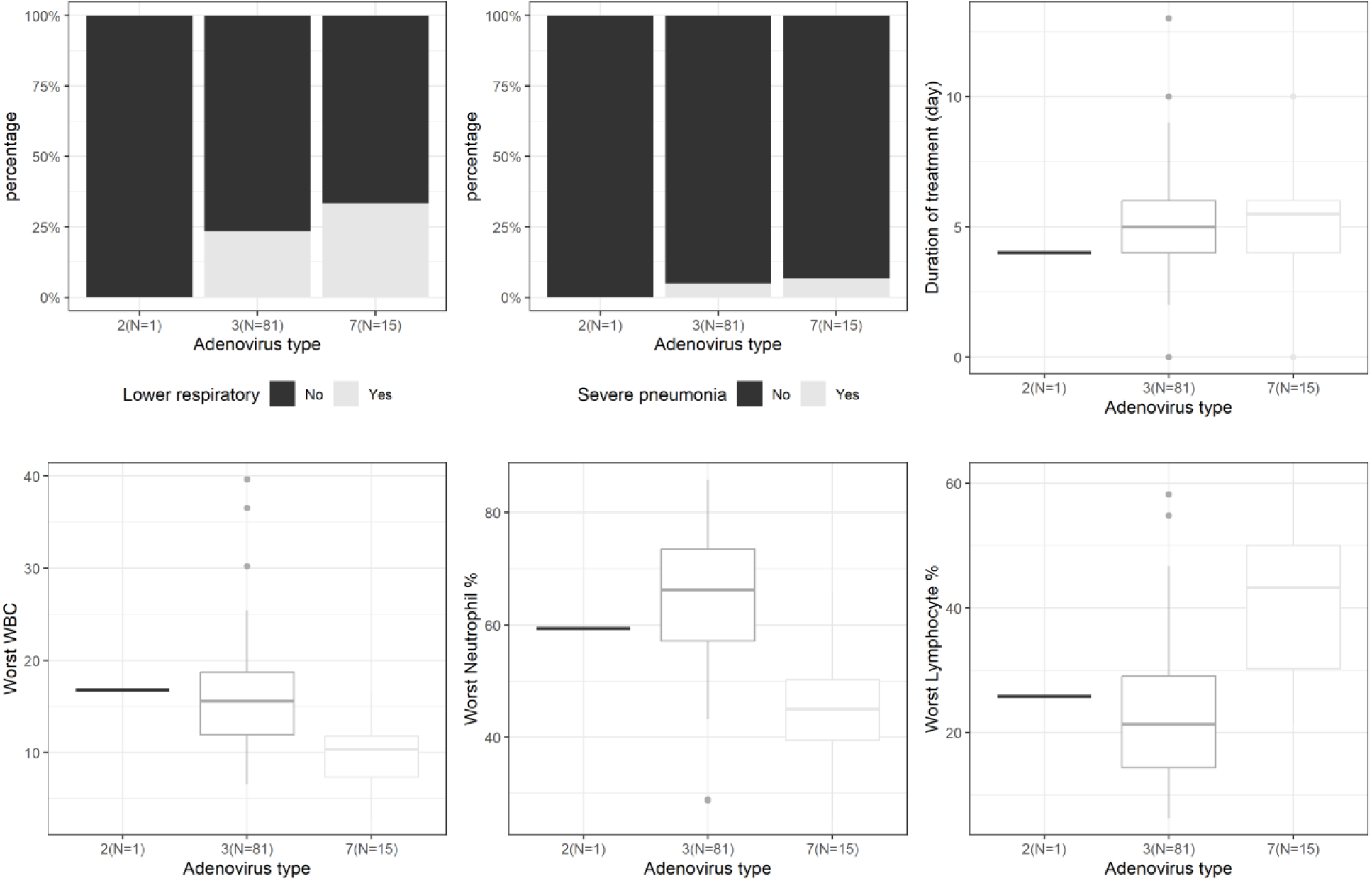
Clinical features by adenovirus types. Upper panel: some clinical features (lower respiratory illnesses (yes/ no), severe pneumonia (yes/ no), duration of treatment) of three groups of patients infected with human Adenovirus (HAdV) type 2 vs. 3 vs. 7. Lower panel: some hematological features (worst white blood cell (WBC) %, worst Neutrophil %, worst lymphocyte %) of three groups of patients infected with human Adenovirus (HAdV) type 2 vs. 3 vs. 7.

## Discussion

This study, to the best of our knowledge is probably the first providing a quick comprehensive snapshot of both clinical features and molecular typing of the respiratory HAdV infection outbreak in children in northern Vietnam in the end of 2022.

The predominant HAdV type in this study was species B type 3 (83%), followed by species B type 7 (16%) and species C type 2 (1%). This is similar to the findings of other studies on circulating or outbreaks of respiratory HAdV infections in children before the COVID-19 pandemic in Asia such as studies on circulating respiratory HAdV in Hong Kong in 2014 (49), Taiwan from 2002 to 2013 (15), Guangzhou China from 2017 to 2019 (27) or a study on febrile respiratory HAdV outbreaks in Hangzhou China in 2011 (17) or a study on successive respiratory HAdV outbreaks in Seoul, Korea from 1990 to 2000 (11). Novel genome types were identified during the outbreaks of lower respiratory tract HAdV infection in Seoul Korea and this 11-year study also suggested a genome type shift between successive outbreaks (11). Therefore, further studies examining the genome types or variants of HAdV may be useful.

There was no significant association between the 3 HAdV types identified and clinical features in this study, except that WBC and neutrophil numbers were higher in those infected with HAdV type 3 than with type 7. The proportion of severe cases seemed to have similar distribution between the more predominant HAdV type 3 and type 7. This is consistent with other previously published studies in which HAdV type 3 and 7 were the most common types among more severe respiratory HAdV infections (10–12,15–19).

Co-infections of HAdV with other respiratory viruses or bacteria stood out to be highly significantly associated with increased severity of respiratory diseases. About 70.8% of those with lower respiratory illnesses and 100% of those with severe pneumonia were co-infected with at least one respiratory viruses or bacteria while the percentage was about 24.7% in those without lower respiratory illnesses. This association remained statistically significant in a multivariate analysis adjusting for other potential risk factors which were either clinically relevant or also statistically associated with lower respiratory illnesses or severe pneumonia. This indicates that the association is both clinically meaningful and statistically robust. This remarkable association suggests that HAdV might be the cause of the outbreak but might not be the main cause of severe cases. Instead, the co-infection of other respiratory viruses or bacteria together with HAdV might increase the severity of the disease. This advocates the advantage of multi- factor respiratory PCR panel(s) for common respiratory viruses and probably also respiratory bacteria together with other microbial tests such as culture for the diagnosis and prognosis of children with respiratory infections. These results may be helpful for the diagnosis, treatment and prognosis of patients in future outbreaks of respiratory HAdV infection.

Although the sample size of this study is limited, it is similar to those of many other previously published studies analysing molecular typing for HAdV outbreaks (5,9,16,49–51). The study samples were from pediatric patients of a private general hospital in Hanoi, Vietnam and molecular typing was performed using samples with Ct for HAdV <30. As such, the study samples may not fully represent the general child population of the outbreak. Additionally, it should be noted that clinical data were collected retrospectively for samples with HadV-positive PCR and thus some clinical data features maybe incomplete, especially for outpatients. Moreover, molecular typing data may not be sufficient to understand the characteristics of HAdV causing the outbreak and thus further study to examine HAdV genome types or variants may be useful.

In summary, this study revealed that HAdV type 3 and type 7 were predominant in the current ongoing outbreak of respiratory HAdV infection in children in northern Vietnam and HAdV type might not be associated with the severity of the diseases. Instead, co-infection of HAdV together with other respiratory viruses or bacteria appeared to be a significant risk factor for lower respiratory tract illnesses and severe pneumonia. These findings advocate the advantages of multi-factor microbial panels for the diagnosis and prognosis of respiratory infections in children. These results may be helpful for the diagnosis, treatment, and prognosis of patients in future outbreaks of respiratory HAdV infection.

## Data Availability

The authors confirm that the data supporting the findings of this study are available within the article and its supplementary materials.

## Notes

### Author Contributions

D.D.N, L.T.T, H.T.T.T., and N.T.H. designed the study as well as drafted the initial manuscript. N.T.H. led the data management and statistical analysis. All authors provided (1) additional contributions to the conception or design of this work or the acquisition, analysis, or interpretation of data for the work; (2) drafting of the manuscript or revising it critically for important intellectual content; and (3) approval of the final version submitted. D.D.N., H.T.T.T., and N.T.H had full access to all the data in the study and take responsibility for the integrity of the data and the accuracy of the data analysis.

